# EEG-Based Frequency Domain Separation of Upward and Downward Movements of the Upper Limb

**DOI:** 10.1101/2023.12.11.23299840

**Authors:** T.V. Ahangama, G.M.K.G.G.B. Gurunayake, I.A. Yalpathwala, J.V. Wijayakulasooriya, T.L. Dassanayake, N. Harischandra, Kwangtaek Kim, R.D.B. Ranaweera

**Author notes:** These authors contributed equally to this work (T.V. Ahangama, G.M.K.G.G.B. Gurunayake).

## Abstract

For a seamless integration of electroencephalography (EEG)-based motor imagery brain–computer interfaces (MI-BCIs), it is vital to be able to classify movements of the same joint. However, a fundamental challenge in classifying the same joint movements arises from the close spatial proximity of the corresponding brain regions. To address this challenge, we explore the feasibility of distinguishing up and down movements specific to the right upper limb using multiple frequency bands combined with a channel averaging method. Six electrodes positioned in close proximity to the motor cortex and two distinct frequency bands: mu (8-12Hz) and beta (12-30Hz) were selected. This isolates and enhances electromagnetic activity in the brain commonly associated with motor and cognitive processing. The results of our study revealed promising outcomes across two classification methods. Utilizing a support vector machine (SVM) classifier, our proposed approach achieved an average accuracy of 59.3% and a k-nearest neighbor(KNN) classifier approach yielded an average accuracy of 61.63% in distinguishing between upward and downward movements of the right arm. These results demonstrate the potential of combining spatially focused EEG acquisition with frequency-specific analysis for improved MI-BCI performance.

## 1 Introduction

Brain-computer interfacing (BCI) is a technology that has the potential to both improve functions in healthy people and also restore usable function in people who are severely affected by a wide range of debilitating neuromuscular illnesses. The primary objective of BCI signal processing is to extract characteristics from obtained brain signals and convert them into logical control instructions for BCI applications^1^. Applications of BCIs based on electroencephalography (EEG) that use motor movements and motor-imagery (MI) data have the potential to be revolutionary in the clinical and entertainment fields. They excel by not necessitating external stimuli, remaining cost-effective, and being entirely non-invasive. Moreover, the discrete movement intention paradigm which utilizes the EEG signals collected before movement onset can be directly used in motor rehabilitation and in navigating through the environment accordingly as it detects movement intention^2^. Such advancements highlight the growing need for deeper understanding and more robust techniques for decoding motor intentions from EEG. However, successfully translating these intentions into reliable control signals remains a challenge due to the poor spatial resolution of surface-recorded EEG and the complexity of EEG signal interpretation, particularly when distinguishing subtle motor actions.

The motor cortex contains unique, spatially separable regions corresponding to specific areas of the human body. This somatotopic organization, often referred to as the motor homunculus, enables distinct brain regions to control specific limbs or muscle groups. Spatial filtering techniques such as common spatial patterns (CSP) are widely used to enhance class-discriminative information by maximizing the variance difference between conditions, especially for binary motor imagery tasks. Consequently, event-related desynchronization (ERD) or event-related synchronization (ERS) induced in corresponding brain areas by different limb movements can be recognized by algorithms that identify distinct spatial activity patterns^3,4^. ERD typically reflects a decrease in mu (8–12 Hz) and beta (13–30 Hz) band power during movement or motor imagery, while ERS reflects a post-movement rebound of oscillatory activity. Therefore, the most widely deployed left and right upper limb, or upper and lower limb movements, serve as the basis for motor imagery-based control systems. Numerous research studies have shown improved accuracy in classifying different limb motions in various applications^5–9^. However, these studies typically focus on movements with clear spatial cortical separation, which may not generalize well to decoding more subtle or fine motor actions in the same limb.

The classification of EEG signals that correspond to the same limb is more challenging than that of separate limbs because the EEG signals that correspond to the same limb movements originate in adjacent areas in the motor cortex with minimal spatial separation. Several studies have shown that brain activity captured by EEG can be used to decode motor intents associated with the same limb activities such as reaching, grasping, finger movements, and complex limb movements^10–15^. These studies show that this spatial proximity leads to overlapping cortical activations and scalp-EEG recordings, which limits the effectiveness of traditional spatial filtering or topography-based classification methods.

However, it is even more challenging to separate the EEG signals induced due to distinct movements done by the same joint (such as flexion and extension of the shoulder), as the differences among spatial activations get even smaller^16^. This becomes a critical issue when developing BCIs for fine motor control, such as in prosthetic fingers, exoskeleton joints, or upper-limb rehabilitation, where distinguishing subtle variations in joint movement is essential. Therefore, in contrast to spatial separation, it is essential to focus more on identifying unique features of the EEG signals triggered by different motor movements carried out by the same joint. This forms the core motivation of our study, which investigates whether fine-grained joint-level distinctions, specifically, upward and downward arm movements, can be detected using targeted signal processing techniques.

Many recent studies continue to apply increasingly complex models in pursuit of marginal gains in accuracy for different limb movements^17–19^, where cortical activations are more easily separable. Additionally, such approaches may introduce unnecessary computational overhead or risk overfitting, particularly in real-time or low-data settings. In contrast, we examine the feasibility of separating upward and downward arm movements in the frequency domain using EEG signals recorded from electrodes positioned over brain regions associated with motor planning and execution. EEG segments both before and after movement onset were analyzed to capture relevant activity. Despite the task complexity, the low computational cost of our model makes it well-suited for potential real-time applications, especially in scenarios with limited data.

The remainder of this manuscript is structured as follows: Section 2 describes the materials and methods, including dataset preparation, electrode selection, signal preprocessing, and feature extraction/classification. Section 3 presents the results, followed by Section 4 with the discussion and directions for future work. Section 5 concludes the study.

## 2 Material and Methods

The complete methodology used in classifying the upward and downward movements performed by the right arm is shown in Figure 1. Comprehensive details of the proposed methodology are provided in the following section.

**Figure 1.**
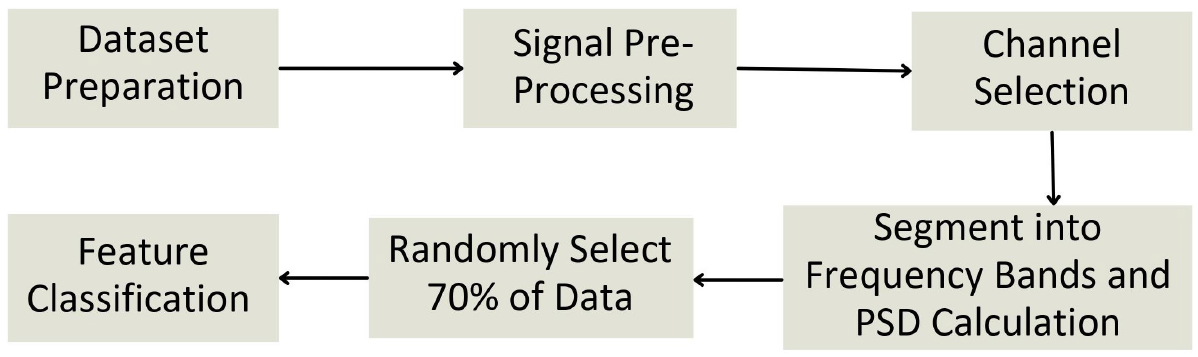
The block diagram illustrating the proposed methodology for identifying the upward and downward movements performed by the same joint.

### 2.1 Data Acquisition and Description

The EEG data for this study was sourced from the “Multimodal signal dataset for 11 intuitive movement tasks from single upper extremity during multiple recording sessions”^20^. This dataset includes 60-channel EEG, 7-channel electromyography (EMG), and 4-channel electro-oculography (EOG) signals recorded at 2500Hz across three days. It was gathered from 25 healthy right-handed subjects and notch filtered at 60Hz to reduce external electrical noise. Electrode positioning followed the international standard 10-20 system^21^. The original experiment involved upper extremity actions, including arm-reaching in six directions, hand-grasping of three objects, and wrist-twisting with two motions performed by the right hand^20^.

Figure 2 illustrates the timing diagram for both real and motor imagery tasks. The original experiment was initiated with a 4-second resting stage, after a gray background and a black cross displayed on the monitor. Subsequently, participants received a visual cue with task instructions for 3 seconds. Following this, participants were presented with text serving as a visual cue indicating whether to execute a motor activity or engage in motor imagery within a 4-second time frame^20^.

**Figure 2.**
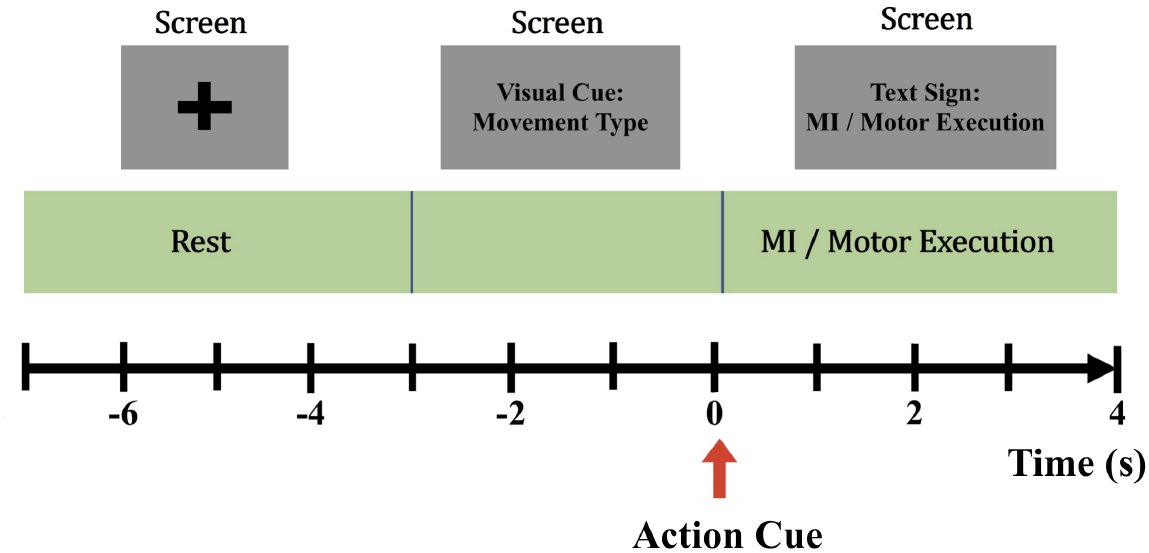
The timing diagram for motor imagery (MI) and motor execution tasks. Each trial begins with a rest period accompanied by a fixation cross (−7s to −3s), followed by a visual cue indicating the movement type (−3s to 0s). At 0s, an action cue prompts the MI or motor execution phase (0s to 4s).

### 2.2 Dataset Preparation

In the original dataset, each subject performed 50 movements from each type of arm reaching real movements randomly. Corresponding EEG (60 channels), EMG (8 channels), and EOG (4 channels) data were recorded simultaneously for all 300 movement epochs^20^. Out of them, data from the EEG channels were selected and subjected to bandpass FIR filtering (0.5-100 Hz) to eliminate high-frequency noise and drifts in all real-reaching movements. Since this study explores only the upward and downward movements, the epochs corresponding to these movements were segregated from all the recorded reaching real movements considering the given trigger points. This separation was done in such a way that each subject had 100 epochs, with 50 epochs per movement type.

### 2.3 Signal Pre-Processing

EEG functions by measuring the integration of all postsynaptic potentials of the populations of neurons over the scalp and has a lower spatial resolution compared to functional Magnetic Resonance Imaging (fMRI), Electrocorticogram (ECoG), etc. Therefore, EEG data are contaminated with artifacts including ECG, eye movements, voluntary muscle activity, and noise from surrounding electronic equipment^22^. Hence, following preprocessing was done on the data set to remove these noise artifacts to improve the SNR.

All the data were resampled at 500 Hz and the artifactual epochs were removed considering the extreme values and abnormal trends. It was verified that no noisy and corrupted channels were present, and then the data were re-referenced with Common Average Referencing (CAR)^23^ as stated in equation 1 to reduce the background noise.

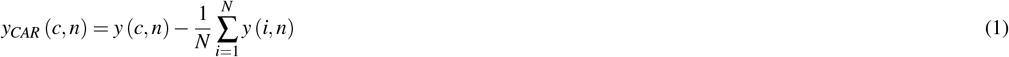

where *y*_*CAR*_(*c, n*) denotes the common averaged potential at channel *c* and sample point *n* which was calculated by subtracting the average potential of total *N* channels at that sample point from EEG potential *y*(*c, n*) at channel *c* and sample point *n*.

Subsequently, independent component analysis (ICA)^24,25^ was performed using the infomax algorithm as it has shown better performance in isolating artifacts^26^. After inspecting the spatial topography, temporal variation, and power spectral density of the isolated components, the independent components identified as artifacts were removed to correct the EEG. Typically, two independent components that contain eye blink artifacts were selected per subject because eye blinks are typically 10 times larger in amplitude compared to ongoing EEG signals, which has a huge impact on the preferred data. This correction minimizes information loss while improving the signal-to-noise ratio (SNR).

### 2.4 Channel Selection

Electrode positioning in the 10-20 system corresponds to different brain regions, and the EEG signals obtained from each electrode represent activity in those specific areas. While using multiple electrodes can help capture a broader range of brain activity, it may also introduce signals unrelated to the joint movements of interest. Since joint movement control is typically localized to a smaller brain region, incorporating signals from widespread electrodes can mix in unrelated activity, potentially distorting features relevant to the task^27^. Therefore, careful spatial localization is essential to ensure that the extracted features accurately reflect motor-related cortical activity. In particular, joint movement–related potentials are not confined to the primary motor cortex alone. These movements may also involve activity originating from the premotor and supplementary motor cortices^28^, emphasizing the need to include only spatially relevant electrodes.

In addition to spatial localization, selecting the appropriate temporal segment of EEG data is equally important. EEG rhythms that discriminate joint movements can be modulated not only during the movement execution phase but also in the period preceding it, which reflects movement intention. This pre-movement activity has been shown to carry valuable information for decoding motor actions^14,29^. Therefore, the inclusion of pre-movement intervals can significantly influence the quality and interpretability of the extracted features.

Hence, this research was conducted by selecting EEG signals from a specific subset of electrodes based on the spatial correspondence between cortical regions and the muscles involved in right upper limb movement^30^. Considering the high probability of capturing relevant neural activity from the motor cortex responsible for controlling the right upper limb^31^, we selected electrodes FC5, FC3, FC1, C5, C3, and C1, as illustrated in Figure 3. The C1, C3, and C5 channels were selected because they are located over the arm area of the motor homunculus of the left primary motor cortex, which is primarily responsible for right-side motor control^32^. FC1, FC3, and FC5 channels were selected as they were placed covering the left half of the pre-motor and the supplementary motor cortices. The supplementary motor cortex is mainly involved with motor planning whereas the pre-motor cortex is responsible for carrying out movement, sequences of movements, and the selection of movements based on sensory information^32^.

**Figure 3.**
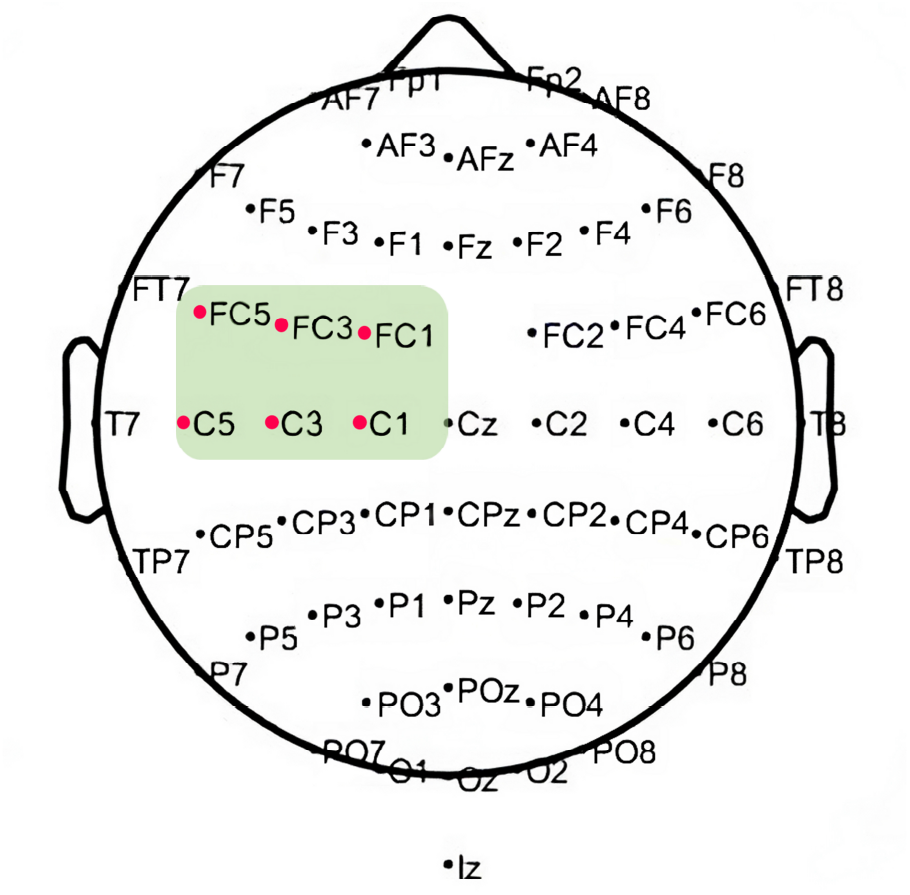
Selected electrodes (highlighted in green) over the left motor cortex for analysis. The six electrodes (FC5, FC3, FC1, C5, C3, C1) were chosen based on their spatial correspondence to motor planning and execution areas related to right upper limb control.

After deriving the event-related potentials (ERP) from the selected epochs, the time segment from –0.5 to 2.5 seconds relative to action cue onset was chosen from these electrodes for further analysis. This ERP-guided selection ensured that the chosen time window captured the neural activity most relevant to motor preparation and execution. As the next step in our approach, a single signal was derived by averaging each epoch across the selected channels, in order to consolidate the effects of signals that were highly correlated with motor movements. This technique was expected to reduce noise and unrelated brain activity, while preserving information from the relevant motor areas.

### 2.5 Feature Extraction

The spontaneous EEG recordings, also known as spontaneous electrical activity, have distinct waveforms that predominate over a wide range of frequencies. Scientifically, there are five frequency bands of spontaneous EEG activity. Each of these frequency bands has a rhythmic activity that is characterized by a specific scalp distribution and biological importance^33^. Among them, mu and beta bands are widely used in MI-based BCI studies due to their correlation with motor activities.

The mu (8-12 Hz) and the Beta (13-30 Hz) Sensory-Motor Rhythms (SMR) have proved to undergo modulations whenever there is a movement of a large body part which are most prominent in EEG signals acquired from C3 and C4 electrodes^34^. This is a phenomenon known as Event-related synchronization (ERS) and Event-Related Desynchronization (ERD) which occur during movement and relaxation respectively. These ERDs and ERSs which lead to short-lasting and circumscribed attenuation of mu and beta rhythms have played an essential role in implementing reliable EEG-based BCI systems across the past three decades. Despite SMR bands not containing kinematic parameter information which has been confirmed by several studies^35,36^, they have been used to distinguish among different motor movements and they provide a reliable feature for BCI.

Consequently, in this study, band-pass filtering was applied to extract signals in the frequency ranges of 8–12 Hz (mu), 12–30 Hz (beta), and 8–30 Hz (combined mu and beta) from the wideband EEG, using a finite impulse response (FIR) filter with a linear phase response. Welch’s power spectral density (PSD) was then computed for each averaged signal using a window length of 1000 sample points (2 seconds) with 500 sample points (1 second) overlap. PSD values within each frequency band for each epoch were considered as features. The process of spectral feature extraction involved several steps, as shown in equations 2, 3, and 4.

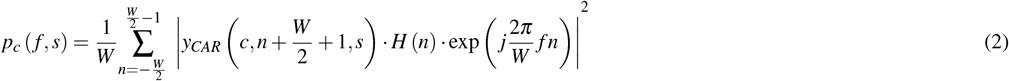

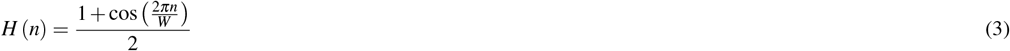

Where *p*_*c*_( *f, s*) denotes the PSD at channel *c* and frequency *f* for segment *s. H*(*n*) is the Hann window of window length *W* set as 1000 sample points. PSD for each trial was normalized by,

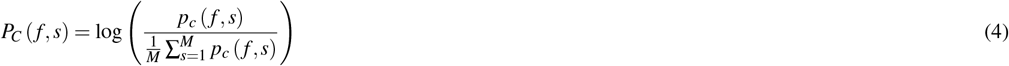

Where *P*_*C*_( *f, s*) is the normalized PSD of each segment with symbols *f, c, s*, and *M* denoting frequency, channel number, segment, and the total number of segments.

### 2.6 Feature Classification

In the EEG signal analysis pipeline, classification methods play a pivotal role, as higher accuracy enables precise motor control of fine movements with high reliability. Despite recent advances in deep learning, traditional machine learning (ML) techniques remain effective, particularly in scenarios with limited data and interpretable features, such as those derived from motor-related EEG rhythms. Among these, support vector machines (SVM), linear discriminant analysis (LDA), and logistic regression have dominated the literature as classification methods for sensory motor rhythm (SMR)-based BCI paradigms, often outperforming deep learning approaches in such contexts^2^.

In this study, the data were randomly split into training and testing sets, with 70% of the trials used for training and 30% for testing. Training data consisted of 35 trials since each subject had 50 epochs for each movement, whereas the remaining epochs were considered as the testing data. Subjects containing less than 35 epochs (n=5) from either up or down movements after the removal of bad epochs were excluded from the analysis. Due to not having enough testing data for several subjects after bad epoch removal, the results were obtained by feeding the 70 training epochs (35 epochs for each movement type) to the classification learner with a K-fold value of 5. To identify the most effective classification algorithm, MATLAB’s classification learner toolbox^37^ was used with automated hyperparameter tuning via Bayesian optimization. This identified SVM and K-Nearest Neighbors (KNN) as the best-performing classifiers for the extracted features. The classification accuracy was evaluated using the Geometric mean (G mean) of the sensitivity(recall) and specificity of the classifiers (equations 5, 6, and 7). Unlike traditional accuracy, which can be inflated by a dominant class, G-mean accounts for both true positive and true negative rates, ensuring that performance reflects the classifier’s ability to distinguish both movement types equally well.

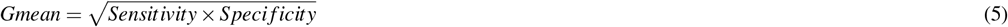

Where,

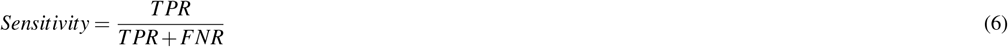

And,

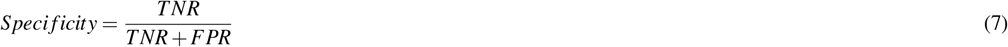

## 3. Results

In the proposed method, three different frequency bands from the measured wideband EEG signals were extracted, intending to identify a particular feature in the frequency domain corresponding to upward and downward movements of the upper limb. Two different classifiers were used to differentiate the extracted features. According to the mean confusion matrices averaged across all 20 subjects (Figures 4 and 5), which present the average sensitivity and specificity values as defined in Equations 6 and 7, both upward and downward movements were classified with an average accuracy of 60% without getting biased for any of the movement types. Table 1 summarizes the sensitivity (recall), specificity, precision, and F1-scores for the SVM and KNN classifiers across all frequency bands.

**Table 1.**
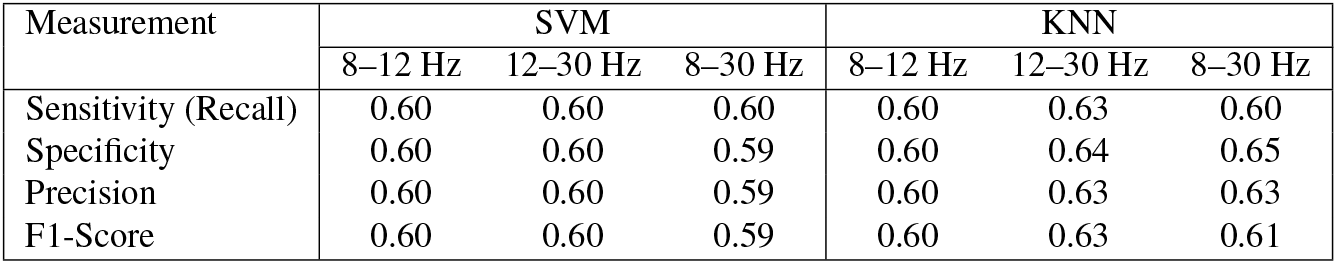
Performance Metrics Across Frequency Bands for SVM and KNN.

| Measurement | SVM |  |  | KNN |  |  |
| --- | --- | --- | --- | --- | --- | --- |
|  | 8–12 Hz | 12–30 Hz | 8–30 Hz | 8–12 Hz | 12–30 Hz | 8–30 Hz |
| Sensitivity (Recall) | 0.60 | 0.60 | 0.60 | 0.60 | 0.63 | 0.60 |
| Specificity | 0.60 | 0.60 | 0.59 | 0.60 | 0.64 | 0.65 |
| Precision | 0.60 | 0.60 | 0.59 | 0.60 | 0.63 | 0.63 |
| F1-Score | 0.60 | 0.60 | 0.59 | 0.60 | 0.63 | 0.61 |

**Figure 4.**
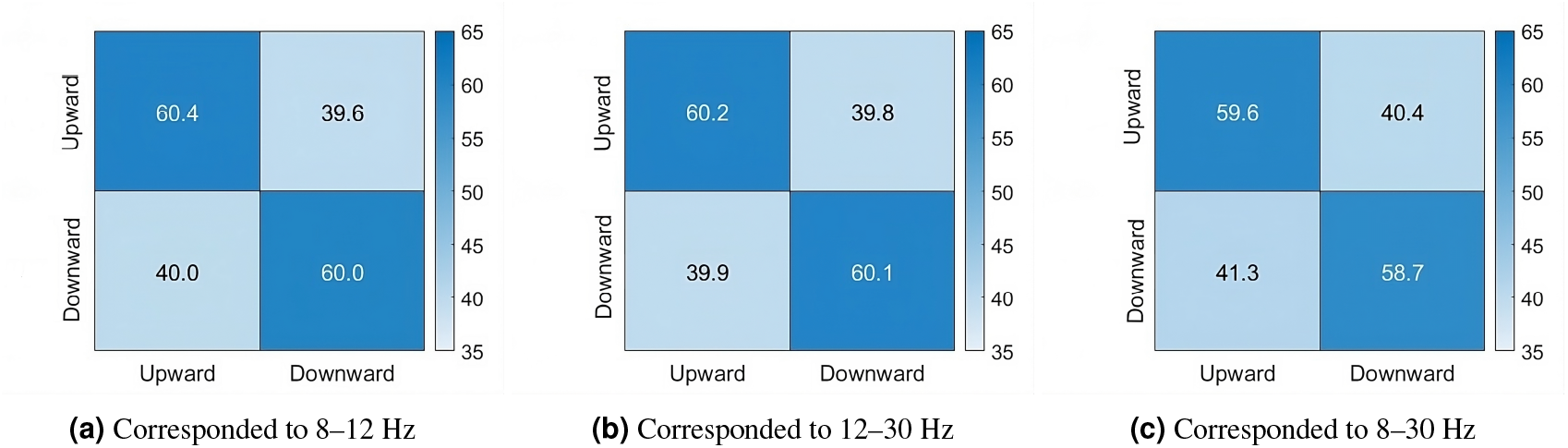
Mean confusion matrices (%) obtained from the SVM classifier. Values shown represent average sensitivity and specificity across subjects (Eq. 6, 7). **(A)** Corresponded to the 8-12Hz. **(B)** Corresponded to the 12-30Hz. **(C)** Corresponded to the 8-30Hz.

**Figure 5.**
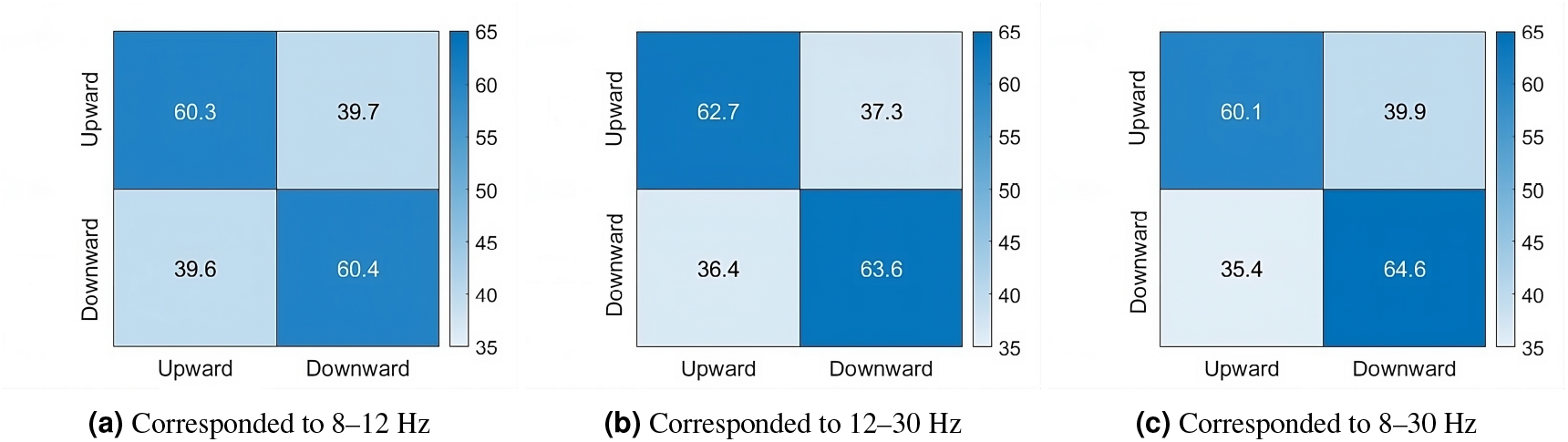
Mean confusion matrices (%) obtained from the KNN classifier. Values shown represent the average sensitivity and specificity across subjects (Eq. 6, 7). **(A)** Corresponded to the 8-12Hz. **(B)** Corresponded to the 12-30Hz. **(C)** Corresponded to the 8-30Hz.

Figures 6 and 7 depict histograms of G-mean values (expressed as percentages) for the SVM and KNN classifiers, respectively, across each selected frequency band for all subjects. Most subjects fall within the 55–65% range. The highest classification accuracy for a single subject was 76% from SVM and 74% from KNN classifiers utilizing the 8-12 Hz band. This suggests some level of inter-subject variability, possibly due to differences in neural activation or engagement during motor imagery. As summarized in Table 2, both the SVM and KNN classification techniques demonstrate averaged Gmean values 5 of approximately 60 ± 1.5% across all three frequency bands, significantly outperforming random chance (p-value *<* 0.00005).

**Table 2.** Overall averaged results of SVM and KNN classifiers across the selected frequency bands. Reported values correspond to the averaged Gmean (Eq. 5) across all subjects, unless otherwise specified. Indicated p-values are for the obtained accuracies with each method compared to a random chance of 0.5.

|  | SVM |  |  | KNN |  |  |
| --- | --- | --- | --- | --- | --- | --- |
|  | 8–12 Hz | 12–30 Hz | 8–30 Hz | 8–12 Hz | 12–30 Hz | 8–30 Hz |
| $\mu$ | 0.60 | 0.60 | 0.59 | 0.60 | 0.63 | 0.62 |
| $\pm\sigma$ | 0.065 | 0.044 | 0.059 | 0.056 | 0.047 | 0.053 |
| p-Value ($\times 10^{-5}$) | 0.435 | 0.006 | 0.288 | 0.048 | 0.0001 | 0.002 |

**Figure 6.**
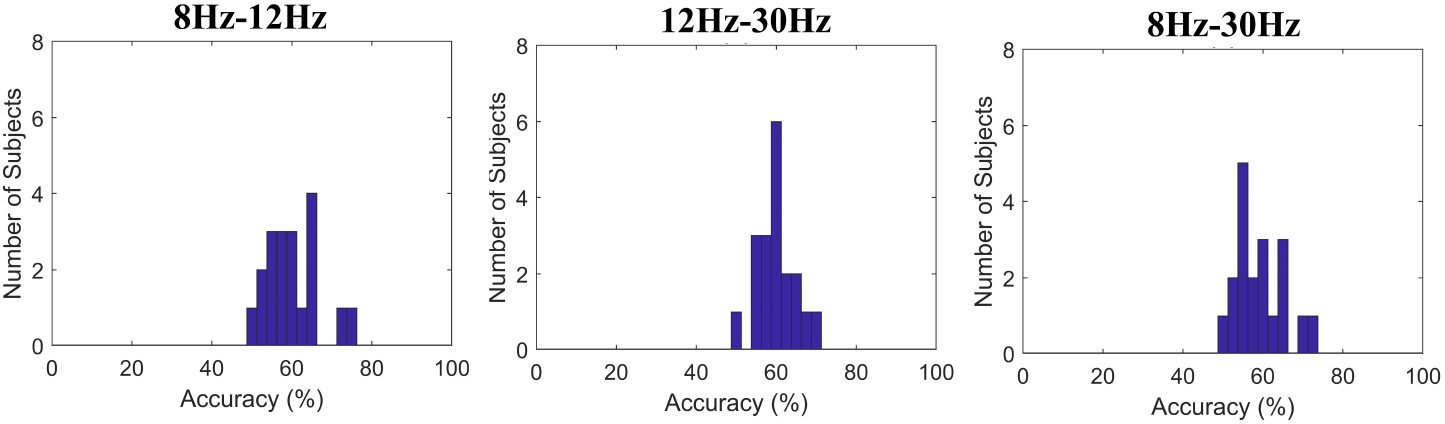
Distribution of subject-wise classification accuracies (Eq: 5) for SVM across SMR frequency bands. The results correspond to mu (8–12 Hz), beta (12–30 Hz), and combined mu–beta (8–30 Hz) bands, respectively.

**Figure 7.**
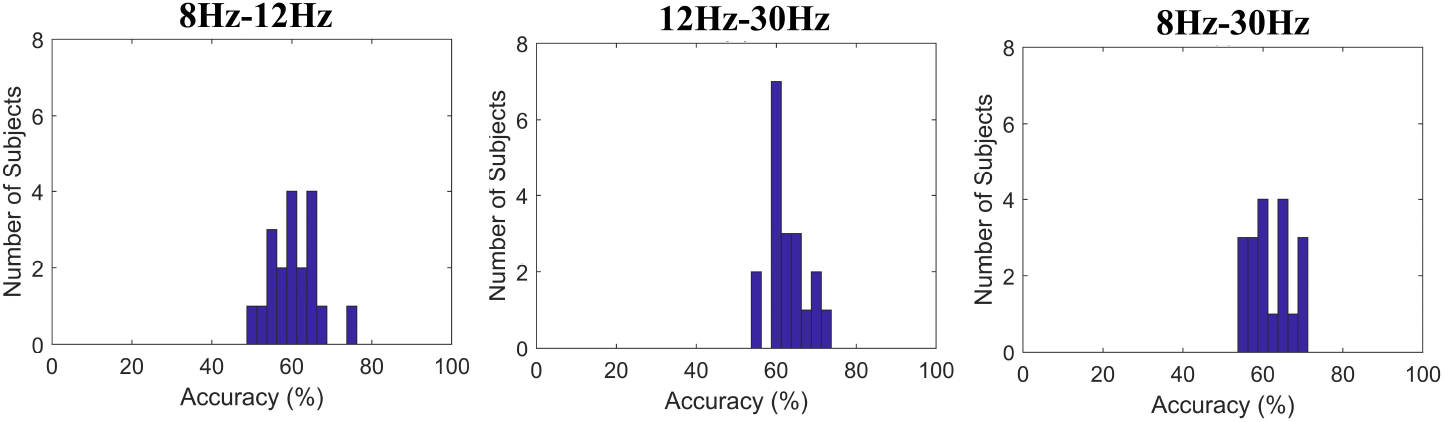
Distribution of subject-wise classification accuracies (Eq: 5) for KNN across SMR frequency bands. The results correspond to mu (8–12 Hz), beta (12–30 Hz), and combined mu–beta (8–30 Hz) bands, respectively.

Figure 8 further highlights the overall performance of selected classifiers. The consistency of results across subjects and frequency bands indicates that the classifiers reliably capture meaningful features, enabling accurate detection of movement direction above chance level using the selected frequency bands. The fact that the classification doesn’t seem biased toward either movement type suggests the features we extracted carry useful and balanced information about both upward and downward motions. The highest individual accuracies around 74–76% in the mu band (8–12 Hz) also hint that this frequency range might play a stronger role in distinguishing movement directions for certain subjects. Overall, even with relatively simple classifiers, the method shows potential for detecting fine motor patterns from just a few EEG channels and frequency bands.

**Figure 8.**
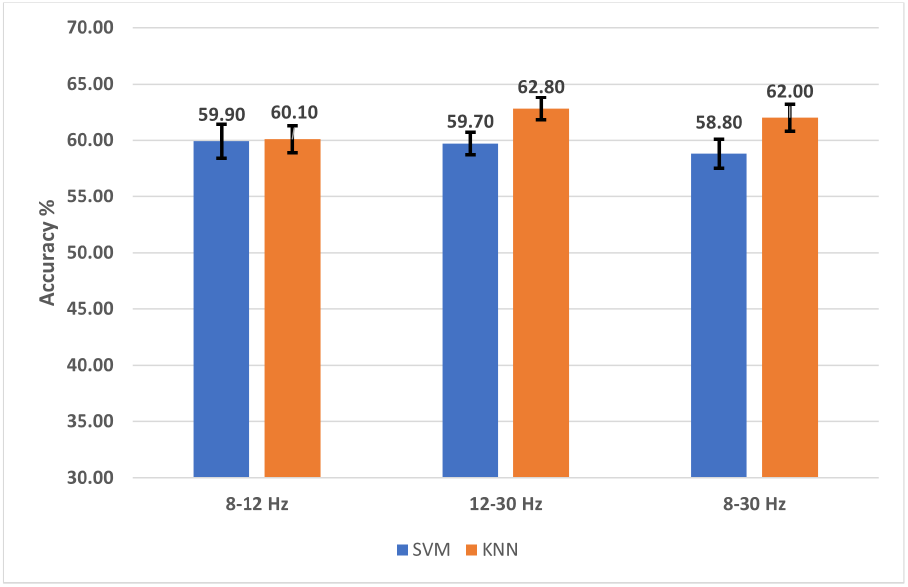
Classification accuracy comparison by both SVM and KNN classifiers. Values shown are averaged across subjects, with percentages reflecting Gmean-based classification performance (Equation 5).

## 4 Discussion

Despite the prevalence of complex neural network–based methods for classifying different limb movements^17–19^, Degirmenci et al. have recently explored a statistical significance–based feature extraction approach for classifying MI tasks across four classes (left hand, right hand, feet, and tongue) using 22 scalp EEG channels^38^.Their method involved extracting features across time, frequency, time–frequency, and non-linear domains, achieving a peak multi-class accuracy of 47.36% and binary-class accuracy of 63.04%. Interestingly, their channel-wise analysis revealed that statistically significant features were concentrated in a subset of motor-related channels, particularly in the mu and beta bands. Nevertheless, classification was performed using all 22 channels, possibly introducing non-motor noise and reducing discriminability—likely contributing to the relatively low performance despite the use of separable tasks.

Considering the separation of the same limb movements in recent studies, five different complex activities executed by the same limb which activate different regions of the brain were differentiated with an average classification accuracy of 94.0 ± 2.7% by^14^ using CSP. However, when the movement types become similar to each other the discrimination becomes difficult as the activation regions draw closer among movements. Xu et al. have obtained about 60% and 40% accuracy in 2-class and 3-class classifications respectively using EEG signals that occurred due to MI-hand, MI-forearm, MI-arm, and rest with 4 different sets of electrodes^39^ while Ma et al. have separated imagery movement of the right hand, right elbow, vs resting with eyes open at an average accuracy of 68.68 ± 2.44%^40^. It is worth mentioning that in some research studies, the reported overall accuracy for multiclass movement classification includes the “rest” category. As a result, the accuracy tends to be higher because distinguishing between movement and rest is generally done with greater accuracy. Several seminal studies have confirmed that kinematic parameters of motor movements (e.g., position, velocity) are embedded in SMR bands below 2Hz^35,41,42^. Using time domain features in low-frequency EEG Ofner et al. have achieved accuracies of 55% (movement vs. movement), 87% (movement vs. rest) for executed movements, and 27% (movement vs. movement), 73% (movement vs. rest) for imaginary movements, elbow flexion, elbow extension, forearm supination, forearm pronation, hand close, and hand open^43^. However, decoding accuracies of kinematic information from low-frequency SMR bands have been reported to be poor^44^ and, the average accuracies of 46.8% in the 5-class scenario and 53.4% in the 4-class scenario obtained from Ma et al. for four different joints and the resting state, using time distributed attention networks show that information within mu and beta frequency ranges in MI task perform better^45^. Achanccaray & Hayashibe et al. have achieved maximum mean accuracies of 78.46 ± 12.50% and 76.72 ± 11.67% for two (flexion/extension) and three (flexion/extension/grasping) class MI tasks respectively using deep learning algorithms^13^. Further, it is evident through results that movements involving different joints are better distinguishable than movements involving the same joints^43^.

This underscores the increased difficulty of classifying movements involving the same joint, making it all the more noteworthy when such movements are accurately distinguished using simpler methods. However, an effective approach to extracting features that yield satisfactory accuracy for discriminating movements involving the same joint remains elusive in current research, to the best of our knowledge. While recent research on dynamic task-incremental learning approaches, such as EEG-DTIL^46^, has achieved 74.92% accuracy in classifying arm up vs.down MI tasks using a deep, multi-stage network with broad learning systems and Fisher-based regularization, our study demonstrates that comparable motor discriminability can be obtained for the same movement pair using a computationally simple, interpretable approach based on Welch’s PSD and traditional classifiers. While EEG-DTIL leverages a high-density 64-channel setup and complex sequential learning strategies, we achieved a peak accuracy of 76% and an average accuracy of 60% using only 6 spatially focused electrodes and handcrafted spectral features in the mu and beta bands.

In comparison to state-of-the-art methods, the findings of our study reveal that, despite the inherent challenge of same-joint classification, carefully selected frequency-domain features from motor-relevant channels can effectively separate movement types without requiring complex architectures or large-scale training data. Using electrodes that span the entire scalp may introduce signals related to unrelated brain processes, such as responses to visual or auditory cues, which can confound the analysis. By focusing only on motor-related electrodes, our approach increases the likelihood that the extracted features reflect motor-related neural activity. Moreover, reducing the number of electrodes to just six channels, related to the primary motor, premotor, and supplementary motor cortices, not only improves reliability but also lowers the computational cost of EEG acquisition.

However, the following limitations of our study can be identified. Due to inter-subject variability, models must be trained separately for each subject, which increases the overall training time and limits the generalizability. While visual inspection allows the removal of artifactual epochs in offline analysis, a real-time BCI system operates under the constraint of processing these potentially problematic epochs without manual intervention. Additionally, the use of non-causal filters in preprocessing is chosen to avoid signal distortion and would need to be replaced with causal filters in real-time systems, which introduces latency.

Future work should explore the impact of electrode selection and time–frequency feature extraction on classification performance. Investigating subject-independent models or transfer learning approaches could improve cross-subject generalization. Incorporating automated artifact detection and rejection algorithms, such as ICA, adaptive filtering, or frequency- and amplitude-based methods may help bridge the gap between offline preprocessing and real-time signal quality requirements. However, recent evidence suggests that removing artifacts may not be essential, and in some cases, it can even be detrimental due to the loss of statistical power if not handled properly^47^. Therefore, the trade-off between preprocessing complexity and real-time performance should be carefully examined when designing cost-effective, real-time BCI systems.

## 5 Conclusions

The lower spatial separation of cortical activations makes it challenging to classify EEG signals elicited by movements involving the same joint. Accordingly, this study aimed to identify discriminative features in the frequency domain associated with such motor activities. During the feature extraction process, the EEG signals induced by up and down right-upper limb movements were averaged across electrodes positioned over motor-relevant cortices. Mu (8–12 Hz), beta (13–30 Hz), and combined (8–30 Hz) frequency bands were then isolated, and Welch’s Power Spectral Density (PSD) was calculated to explore their discriminative potential. Classification was performed using traditional machine learning algorithms, specifically SVM and KNN, to preserve computational simplicity.

Our findings demonstrate that frequency-domain features over a small set of motor-relevant channels can distinguish same-joint movements with promising accuracy. This supports the viability of simple, interpretable models in scenarios where complex deep learning architectures may not be feasible. The results contribute to ongoing efforts in designing cost-effective, real-time BCI systems, particularly for applications involving subtle motor control. Future work could focus on expanding the model’s generalizability across subjects, optimizing real-time implementation with causal filters, and further investigating the role of electrode selection and time–frequency features in improving classification accuracy for same-joint movements.

## Author Contributions

T.V.A. and G.M.K.G.G.B.G. contributed equally to the study. They were responsible for conceptualization, methodology, software, formal analysis, investigation, visualization, data curation, validation, and writing – original draft, review & editing. I.A.Y. contributed to data curation, validation and writing – original draft. R.D.B.R. supervised the study and contributed to conceptualization, writing – review & editing and project administration. T.L.D., J.V.W., N.H., and K.K. were involved in writing – review & editing and provided intellectual input during manuscript development. All authors approved the final version of the manuscript and agreed to be accountable for its content.

## Data availability statement

The data that support the findings of this study are openly available in the GigaDB repository, at http://dx.doi.org/10.5524/100788

